# COMPARISON OF SARS-COV-2 WUHAN AND ALPHA VARIANTS: CLINICAL AND LABORATORY HIGHLIGHTS

**DOI:** 10.1101/2022.05.17.22275188

**Authors:** Demet Timur, Uğur Demirpek, Engin Ertek, Özlem Çetinkaya Aydın, Talha Karabıyık, Hüseyin Kayadibi

## Abstract

Since December 2019, after the declaration of new cases regarding novel coronavirus disease, many variants have emerged as a consequence of the viral evolution. Though the SARS-CoV-2 variants have been studied for molecular basis, the clinical and pathologic disparities of them have been understood inadequately. The aim of this research was to figure out the differences between the SARS-CoV-2 Alpha (B1.1.7) variant and the classical Wuhan groups on the clinical basis and laboratory results of the COVID-19 patients who had positive PCR test.The study was done retrospectively inclusive of epidemiological, laboratory data and clinical symptoms of patients who were admitted to the emergency service between February 15 and March 15, 2021 and had positive COVID-19 PCR test results. Though there was no statistically significant difference in symptoms between SARS-CoV-2 Alpha variant and classical variant (Wuhan type) groups; C-reactive protein (CRP), lymphocyte and leukocyte counts were statistically significantly higher in the Wuhan type group; prothrombin time (PT), International Normalized Ratio (INR) and serum creatinine values were statistically significantly higher in the Alpha group. Studies such as ours that investigate both the clinical features and laboratory data of SARS-CoV-2 variants will close the knowledge gaps, so better decisions may be made by health policy makers. Additional studies in this area will increase the understanding of the topic.

## INTRODUCTION

The globe had burst into Severe Acute Respiratory Syndrome Coronavirus 2 (SARS-CoV-2) disease in December 2019 and, soon it became a pandemic. For the sake of combat against disease, it is necessary to fathom the clinical pathologic state and analysis of translational data. The Coronavirus disease 2019 was observed in wider clinical spectrum in adult patients [1]. This spectrum could lead to different clinical pictures ranging from asymptomatic ones to the patients requiring intensive care hospitalization and even resulting in death. In March 2022, the number of deaths from SARS-CoV-2 infection were approximately six millions globally, while the number of cases were approximately 462 millions. During COVID-19 pandemic, different SARS-CoV-2 genetic variants have emerged globally and studies have focused on genotypic properties of the emerging strains. Centers for Disease Control and Prevention (CDC) have identified Alpha (B.1.1.7), Beta (B.1.351), Gamma (P.1), Epsilon (B.1.427, B.1.429), Eta (B.1.525)), Iota (B.1.526), Kappa (B.1.617.1), Delta (B.1.617.2), B.1.617.3, Mu (B.1.621, B.621.1), Zeta (P.2), Omicron (B.1.1.529) as different genetic variants [2]. Alpha variant was derived from SARS-CoV-2 20B/GR B.1.1.7 strain and contains several mutations including N501Y (conversion of asparagine in S gene at codon 501 to tyrosine) and the 69-70 deletion (deletion of codons at positions 69 and 70 in the S gene) [3, 4]. Understanding of new genetic variants may contribute to better diagnostics, also to the comprehension of both unpredictable increase in contagiousness and the disease severity. Moreover, it has been hypothesized that weak antibodies resulting from previous infection or vaccination may cause treatment unresponsiveness [3, 5]. Alpha variant (B.1.1.7) was first detected in the UK in September 2020 and spread to many countries [6, 7]. After the Alpha variant was also seen in Turkey, it quickly became the dominant strain [8].

Although routine laboratory tests are not specific to the disease, their contribution to the evaluation of severity and progress is pivotal. Because it is imperative to understand concomitant complications and the situation of the organs involved. Based on this, the epidemiological and laboratory data of Alpha and Wuhan type SARS-CoV2 variants were investigated within the scope of the COVID-19 pandemic in our hospital.

## METHODS

The study was single-centered. Demographic data of the patients were obtained retrospectively from the hospital database. Approval for this study was obtained from the Clinical Research Ethics Committee (02.06.2021, Decision no: 2021-10/14). This study was carried out according to the principles of Declaration of Helsinki of human rights. This study was approved by the Turkish Ministry of Health Scientific studies council (2021-03-21T18_47_42).

### Patients’ characteristics

This study included the patient population (n = 379) who visited the emergency service for the suspicion of COVID-19 infection and were diagnosed as COVID-19 by PCR tests. Of our patients diagnosed with COVID-19, 190 were infected with Wuhan type and 189 with the alpha variant. The epidemiological characteristics of the different patients’ groups were similar.

### Laboratory analysis

Samples taken from patients via combined oropharyngeal and nasopharyngeal swabs were stored at 4°C in viral transport medium. After nucleic acid extraction, Bio-Speedy SARS-CoV-2 Variant Plus kit (Bioeksen, Turkey) was used in Rotor-Gene Q device (Qiagen, Germany) for PCR detection. Variant analyses were performed simultaneously with RT-PCR. RNA-Sequencing of the variants were not performed.

Blood urea nitrogen (BUN), creatinine, aspartate aminotransferase (AST) and alanine aminotransferase (ALT) and C-reactive protein (CRP) were measured in a Cobas C 702 (Roche Diagnostics, Germany) analyzer from the sera of the patients. Ferritin, creatine kinase MB (CK-MB), and troponin T levels were measured with Cobas E 801 (Roche Diagnostics, Germany) analyzers. Prothrombin time (PT), activated partial thromboplastin time (aPTT) and D-dimer levels were measured with Cobas T 711 (Roche Diagnostics, Germany) analyzer. Complete blood count was analyzed on Sysmex XN-9100 hematology analyzer (Sysmex Corporation, Japan).

#### Statistical analyses

R-based Jamovi (Sydney, Australia) 1.6.23 was used for statistical analysis. Shapiro-Wilk test was used to determine the conformity of variables to a normal distribution. Student’s *t*-test was used to compare normally distributed parameters between the groups, and Mann-Whitney U test was used to compare non-normal distributed parameters between groups. Chi-square test was performed to investigate whether there was a difference between categorical variables.

## RESULTS

The study included 379 patients who had come to the emergency department between February 15 and March 15, 2021 and had positive COVID-19 PCR test results. 190 (50%) of the patients were Wuhan type (96 Male / 94 Female) and 189 (50%) were alpha variants (96 Male / 93 Female).

The laboratory test results of the patients are shown in Table 1. Leukocyte count (*P*= 0.024), lymphocyte count (*P*= 0.018), PT (*P*= 0.014), INR (*P*= 0.023) and CRP level (*P*= 0.049) were found to be statistically different between the groups. Leukocyte count (*P*= 0.012), lymphocyte count (*P*= 0.009) and CRP (*P*= 0.025) were found to be statistically significantly higher in the Wuhan type group. PT (*P*= 0.007), International normalized ratio (INR) (*P*= 0.011) and creatinine (*P*= 0.042) were found to be statistically significantly higher in the alpha group. When the symptoms were compared between the groups, no statistically significant differences were found for any of the symptoms.

**TABLE 1.** SUMMARY of SYMPTOMS AND LABORATORY TEST RESULTS

| **Variables** | **Wuhan type, n= 190** | **Alpha variant, n= 189** | **P** |
| --- | --- | --- | --- |
| Outpatient, N (%) | 140 (73.7) | 126 (66.7) |  |
| Inpatient, N (%) | 42 (22.1) | 48 (25.4) |  |
| ICU, N (%) | 8 (4.2) | 15 (7.9) |  |
| Gender, Female/Male, n (%) | 94 (49.5) / 96 (50.5) | 93 (49.2) / 96 (50.8) | 0.958 |
| Age, year | 45.5 (24) | 45 (20) | 0.744 |
| WBC ($10^3/\mu\text{L}$), Median (IQR) | 6.22 (2.35) | 5.76 (2.03) | **0.024** |
| Neutrophil ($10^3/\mu\text{L}$), Median (IQR) | 3.69 (2.24) | 3.46 (2.03) | 0.231 |
| Lymphocyte ($10^3/\mu\text{L}$), Median (IQR) | 1.57 (0.85) | 1.46 (0.90) | **0.018** |
| NLR, Median (IQR) | 2.22 (1.78) | 2.15 (2.34) | 0.599 |
| Haemoglobin (g/dL), Median (IQR) | 13.9 (2.48) | 14.2 (2.30) | 0.351 |
| Platelet ($10^3/\mu\text{L}$), Median (IQR) | 212.5 (87.8) | 231 (81) | 0.119 |
| MPV (fL), Median (IQR) | 10.4 (1.3) | 10.2 (1.2) | 0.148 |
| PT (s), Median (IQR) | 8.585 (0.68) | 8.78 (0.81) | **0.014** |
| INR, Median (IQR) | 0.97 (0.07) | 0.99 (0.09) | **0.023** |
| aPTT (s), Median (IQR) | 28.3 (4.7) | 28.9 (4.7) | 0.314 |
| D-Dimer ($\mu\text{g/mL}$ FEU), Median (IQR) | 0.28 (0.3075) | 0.28 (0.27) | 0.897 |
| BUN (mg/dL), Median (IQR) | 11.5 (5.85) | 11.9 (5.70) | 0.521 |
| Creatinine(mg/dL), Median (IQR) | 0.820 (0.2875) | 0.890 (0.35) | 0.084 |
| AST (U/L), Median (IQR) | 20 (11.75) | 21 (13) | 0.223 |
| ALT (U/L), Median (IQR) | 21 (14.75) | 21 (18) | 0.229 |
| Ferritin ($\mu\text{g/L}$), Median (IQR) | 112.5 (162.5) | 110 (212) | 0.676 |
| CK-MB ($\mu\text{g/L}$), Median (IQR) | 1.085 (1.22) | 1.13 (1.08) | 0.712 |
| Troponin T (ng/L), Median (IQR) | 4.25 (3.975) | 4.30 (3.6) | 0.715 |
| CRP (mg/L), Median (IQR) | 7.80 (16.15) | 5.10 (10.50) | **0.049** |
| Fever, n (%) | 54 (28.42) | 51 (26.98) | 0.755 |
| Fatigue, N (%) | 71 (37.37) | 71 (37.57) | 0.968 |
| Cough, N (%) | 110 (57.89) | 116 (61.38) | 0.490 |
| Dyspnea, N (%) | 36 (18.95) | 37 (19.58) | 0.877 |
| Anosmia & Ageusia, N (%) | 23 (12.11) | 15 (7.94) | 0.177 |
| Diarrhea, N (%) | 6 (3.16) | 5 (2.65) | 0.766 |
| Headache, N (%) | 34 (17.89) | 34 (17.99) | 0.981 |
| Arthralgia, N (%) | 43 (22.63) | 43 (22.75) | 0.978 |
| Myalgia, N (%) | 22 (11.58) | 18 (9.52) | 0.515 |
| Sore throat, N (%) | 40 (21.05) | 35 (18.52) | 0.536 |
| Death, N (%) | 6 (3.16) | 8 (4.23) | 0.579 |

WBC: White Blood Cell, NLR: Neutrophil Lymphocyte Ratio, MPV: Mean Platelet Volume, PT: Prothrombin Time, INR: International Normalization Ratio, aPTT: activated partial thromboplastin time, BUN: Blood Urea Nitrogen, AST: Aspartate Transaminase, ALT: Alanine Transaminase, CK-MB: Creatine kinase-myoglobin binding, CRP: C-Reactive Protein.

In this study, when patients with COVID-19 infection were evaluated in terms of mortality, eight patients out of 189 patients with alpha variant and six patients out of 190 patients with Wuhan type had died. There were no statistically significant differences between alpha variant and Wuhan type for the death rates. On behalf of the symptoms encountered in those patients, no significant differences were found between the two groups of this study.

## DISCUSSION

As the new SARS-CoV-2 variants emerge due to the mutations, new variants affect the clinical picture and laboratory data. In our study, we compared the clinical and laboratory data of patients with SARS-CoV-2 alpha variant and Wuhan type.

Symptoms such as fever, cough, weakness, shortness of breath, loss of taste and smell, which are frequently encountered in SARS-CoV-2 infection, are non-specific symptoms. In a study of Bhatraju et al., conducted at ICU, the most common symptoms of COVID-19 patients were cough, shortness of breath, and fever, as well as were observed in 50% of these patients’ at the time of admissions to the hospital [9]. In a study conducted with 1099 patient data in China, when the duration of hospital stay is included, the most common symptom is fever, while the second symptom is cough [10]. Sahin et al., figured out that fever and cough were the most common symptoms [11]. In our study, the most common symptoms were cough, malaise and fever. In another study, Alp et al., found that the clinical presentations of both confirmed and probable cases could vary in a wide spectrum [12]. We concluded that the abundance of clinical spectra could be linked to the presence of different variants (VOC).

In a survey of patients infected with the alpha variant; cough, fatigue, sore throat, myalgia, and a history of fever within 7 days prior to the test were more common, while loss of taste and smell (anosmia and ageusia) were found less [13]. Graham et al., on the other hand, showed that there is no direct relationship between symptoms and alpha variant, as we found [14]. Tsai et al., studied the analysis of deaths occurring in the alpha variant infected 636 patients, the mortality rate of symptomatic patients was low; however, elder patients with no symptoms had higher mortality rate [15]. In our study, no statistically significant differences was found when we compared groups in terms of mortality and symptoms.

In different studies, statistically significantly higher rates of hospitalization, admission to intensive care units, and death were found in the alpha variant compared to the classical variant [16, 17]. Graham et al., indicated that also our research noticed, the rates of asymptomatic cases and hospital admissions were not altered by alpha variant [14].

In COVID-19 infection, lymphopenia, leukocytosis, hypoalbuminemia, neutrophilia, thrombocytopenia and increased troponin, creatinine, AST, ALT, CRP, PT, aPTT, D-dimer levels were observed [18, 19]. In our study, PT and creatinine values were found to be statistically higher in the alpha variant group.

In a study conducted by Guan et al. with the data of 1099 patients, 83% lymphocytopenia, 36% thrombocytopenia and 34% leukopenia were found at the time of admission. While CRP elevation was the most common result; ALT, AST, CK, and D-dimer level elevations were observed less frequently [10]. In favor of inauguration to predict disease progression in COVID-19 at early disease stages, e.g. at hospital admission, the value of biomarkers is imperative. The results have the potential to be used to improve the management of COVID-19 patients for identification of high-risk groups and pertinent handling of healthcare resources in the pandemic. Ozkarafakılı et al., found that there was no association between the human blood groups and laboratory results. In our study, human blood groups were not evaluated [20]. While the WBC counts were higher in severe COVID-19 infections, the lymphocyte counts were statistically significantly lower. A high neutrophil/lymphocyte ratio indicates critical illness and poor prognosis [21, 22].

In addition to the increase in the N/L ratio (NLR), many studies also indicate increases in CRP and D-dimer levels, supporting the severity of the COVID-19 infection [23, 24]. In our study, CRP levels were higher in the Wuhan type group. When the two groups were compared in terms of symptoms, no statistically significant differences were found. In a study conducted by Song et al.; CRP, CK, D-dimer levels were found to be statistically significantly higher in the alpha variant patients’ group while there were no statistically significant differences between the two groups in terms of WBC, neutrophil, platelet, lymphocyte, ALT and AST values [25]. Dagcioglu et al., found that the lymphocytes counts were higher in the alpha variant patients’ group; whereas WBC count, neutrophil count, NLR and ferritin values were found to be lower [26]. In another study with 158 patients, Vassallo et al. found the platelet count to be higher in the alpha variant patient group, while the CRP, D-dimer and NLR were not statistically significantly different between the two groups. The alpha variant was found to have a four-fold higher risk of death and hospitalization in intensive care [27]. In our study, CRP levels, lymphocyte and leukocyte counts were found to be statistically significantly higher in the Wuhan type patient group. These results showed statistically significant differences between the alpha variant and the Wuhan type variant groups.

## CONCLUSION

In this study, no differences were found in terms of symptoms in both variant groups. When laboratory data were examined; CRP levels, lymphocyte and leukocyte counts were found to be statistically significantly higher in the Wuhan type group. PT and creatinine were found to be statistically significantly higher in the alpha variant group. In our study, however, no statistically significant difference was found between the variant groups in terms of mortality and all evaluated symptoms. As mentioned in this study, the increase in the number of studies explaining the clinical and laboratory features of variants could fill in the lack of information on this issue. Nevertheless there were limitations on behalf of this retrospective study, such as low number of patients, being single-centered and absence of RNA-sequencing data. However further studies may help to solve the iceberg illusion.

Timur D, Demirpek U contributed to the conception and design of the study. Ertek E, Aydın OC were involved in clinical evaluation. Demirpek U, Karabiyik T, Timur D interpreted the results. Karabiyik T, Kayadibi H performed of statistical analysis. Timur D, Demirpek U, Karabiyik T, Kayadibi H. drafted the manuscript. Demirpek U supervised the study.

## Data Availability

All data produced in the present study are available upon reasonable request to the authors

## Research funding: None declared

Author Contributions: All of the authors declare that they have participated equally and approved the final version of the manuscript.

## Ethical Consent

Ethical council approval was obtained from local authorities. (02.06.2021, Decision no: 2021-10/14). This study was carried out according to the principles of the Declaration of Helsinki. This study was approved by the Turkish Ministry of Health Scientific studies council (2021-03-21T18_47_42).

## Informed Consent

The authors declare that this study is done retrospectively, no informed consent form used for this study.

